# Modelling malaria routine surveillance data to inform seasonal malaria chemoprevention strategy in Moissala, Southern Chad

**DOI:** 10.64898/2026.03.23.26349112

**Authors:** Nicholas Putney, Jessica Sayyad-Hilario, Israel Ukawuba, Francesco Grandesso, Saschveen Singh, Prince Djuma Safari, Emilie Pothin, Beatrice Filippini, Elkoussing Djovouna, Mahamat Saleh Issakha Diar, Clara Champagne, Anton Camacho

**Author notes:** equal contribution (Clara Champagne, Anton Camacho).

## Abstract

**Background:** Seasonal malaria chemoprevention (SMC) is a malaria intervention in which antimalarial drugs are administered monthly to children under 5 years of age during the high-transmission season. In the district of Moissala in southern Chad, SMC has been implemented since 2013, with an interruption in 2019, resumption in 2020, and expansion to five rounds of treatment in 2021. Recent World Health Organization (WHO) guidelines allow countries to adapt the timing and number of SMC rounds to local transmission patterns, creating a need to identify optimal strategies for each setting. In this study, we used mathematical modeling for three primary purposes: 1) to estimate the effectiveness of SMC in Moissala from 2018 to 2023, 2) to assess the impact of changes to SMC strategies since 2018, and 3) to determine the optimal SMC strategy in Moissala.

**Methods and findings:** We adapted a compartmental, climate-informed malaria transmission model to represent malaria dynamics in the presence of SMC. The model incorporates temperature and rainfall data to capture how climate variability influences malaria transmission over time. It was calibrated to routine surveillance data on malaria cases in children under five years old from 2018 to 2023. Using the calibrated model, we simulated malaria cases under alternative scenarios, including the absence of SMC and variations in the number and timing of SMC rounds. These simulations were then used to estimate the overall effectiveness of SMC, assess the impact of past changes in SMC strategies, and identify the optimal strategy in Moissala. Between 2018 and 2023, SMC reduced malaria cases in children under five by 26% (95% credible interval: 21%, 31%) relative to a scenario without SMC, corresponding to an average of approximately 14400 cases averted each year. The interruption of SMC in 2019 led to an estimated increase of 13600 cases (95% credible interval: 11200, 15800), representing a 31% rise during the high-transmission season. Expanding from four to five SMC rounds in 2021 reduced cases by 7% relative to a four-round schedule, while starting the five-round schedule earlier in June rather than July led to an additional 5% reduction. Overall, the most effective strategy from 2018 to 2023 was a five-round schedule beginning in mid-June.

**Conclusions:** Seasonal malaria chemoprevention has substantially reduced malaria incidence among children under five in Moissala. The currently implemented strategy of five rounds of SMC starting in June was estimated to achieve the greatest reduction in cases over the study period. Climate-informed modelling and open-source software can support timely decision-making across settings under changing climate and transmission conditions.

## Introduction

Malaria remains a leading cause of morbidity and mortality in sub-Saharan Africa, with children under 5 years of age bearing a disproportionate share of the burden. In areas where transmission is highly seasonal, seasonal malaria chemoprevention (SMC), consisting of monthly administration of sulfadoxine-pyrimethamine plus amodiaquine to children aged 3 to 59 months during the high-risk months, has consistently reduced clinical malaria and severe disease in randomized trials and programmatic evaluations, and has been recommended by the World Health Organization (WHO) since 2013 for the Sahel sub-region [1–5]. Building on this evidence base, SMC has been scaled up across West and Central Africa and now reaches tens of millions of children annually [4, 5]. In 2022, WHO consolidated its malaria guidelines and explicitly encouraged tailoring SMC to local epidemiology, including the timing and number of rounds, to reflect variation in the onset and duration of peak transmission across settings [6].

Chad exemplifies the programmatic challenge of aligning SMC with heterogeneous climatic regimes and transmission dynamics. The country comprises three broad geo-climatic zones (Saharan, Sahelian, and Sudanian) with increasing rainfall and lengthening rainy seasons toward the south. Malaria is the foremost cause of outpatient consultations, hospitalizations and hospital deaths nationally, and the burden is especially intense in the southern Sudanian zone which experiences a longer transmission season [7]. The Moissala health district in Mandoul Province is situated in this southern belt and is characterized by a sustained rainy season typically spanning May to October and a marked surge in pediatric malaria from June onward. In 2022, following updated WHO guidance, the National Malaria Control Programme (Programme National de Lutte contre le Paludisme, PNLP) conducted nationwide analyses of malaria seasonality and recommended implementing five SMC rounds in Moissala [8].

Médecins Sans Frontières (MSF) has supported malaria case management in Moissala since 2010, including 23 health centres and a pediatric malaria unit, and has coordinated SMC implementation with national and local authorities since 2013. From 2013 to 2018, four monthly SMC rounds were delivered annually, typically from July to October, in line with WHO’s early operational guidance for Sahelian districts. In 2019, SMC was suspended in Moissala after the district was deemed ineligible under earlier criteria; this interruption coincided with a pronounced increase in reported malaria cases, prompting PNLP to re-authorize SMC in 2020. In response to the longer southern transmission season, a fifth round was added in 2021. Following Epicentre/MSF internal analyses indicating earlier onset of transmission in Moissala, the first round was advanced to June in 2023. Household coverage surveys conducted by Epicentre/MSF around these campaigns consistently documented high uptake (generally 80 − 90%).

Despite the policy shift toward flexible, context-specific SMC, evidence gaps persist on how best to time and scale-up SMC in areas with longer high-transmission seasons, and on the retrospective public-health impact of SMC under routine program conditions. Straightforward year-to-year comparisons are often misleading because malaria incidence is strongly modulated by inter-annual variability in rainfall and temperature, which influence vector breeding, survival, and parasite development [9–14]. Routine surveillance data, while indispensable for decision-making, therefore require analytic frameworks that control for climatic confounding to estimate intervention effects credibly and to compare alternative deployment scenarios [9, 15–17]. Mechanistic, climate-informed transmission models calibrated to local routine data offer a principled approach to this challenge: they can decompose the observed time series into contributions from climate-driven transmission and programmatic interventions, and then simulate counterfactual strategies that would be difficult or impossible to evaluate prospectively [9, 18].

Here, we adapted an existing climate-informed compartmental model of malaria transmission calibrated on data from 2018 to 2023. Using this model, we estimate the effectiveness of SMC, assess the impact of past changes in implementation, and identify the optimal SMC strategy in Moissala, by analyzing counterfactual scenarios that vary the start month and number of rounds, two parameters highlighted in recent WHO guidance [6]. In addressing the above, the study provides operational recommendations intended specifically for the Moissala district and its implementing partners (MSF and the PNLP). The modelling framework is also made available as an open-source R package and can be adapted to other districts or countries to optimize the timing and number of SMC rounds using local surveillance and climate data.

## Data

### Incidence data

Weekly malaria case data have been collected by MSF in the district of Moissala since 2013. Because the data collection tool changed in 2018, the analysis was restricted to the period from 2018 to 2023. We used the number of patients seeking care at any health facility in the Moissala or at Moissala Hospital and diagnosed with malaria. For this analysis, we focused on malaria cases among children under five years of age, the population targeted for SMC, and most affected by malaria.

### SMC coverage data

Household surveys were conducted annually by Epicentre from 2014 and 2023 to estimate SMC coverage in the district of Moissala. During SMC campaigns, children receive antimalarial drugs, and their parents are given an SMC card documenting that the child has received the intervention. At the time of the household surveys, some parents may no longer have this card or may never have received it. Therefore, two SMC coverage indicators were reported: one based solely on presentation of the SMC card, and another that also includes children whose parents verbally confirmed that the child received SMC, regardless of card availability. For our analysis, we used the latter indicator for the 2018 to 2023 period. As no survey took place in 2018, coverage estimates from the 2017 survey were used. In 2019, SMC implementation was suspended, and consequently no survey was conducted.

### Climate data

Daily rainfall data were obtained from CHIRPS, which merges station and satellite observations [19]. Monthly temperature estimates were sourced from the ERA5-Land dataset [20]. For rainfall, monthly cumulative values were first computed, a smoothing spline was fitted, and daily smoothed estimates were derived and standardized (mean=0, variance=1). For temperature, smoothing splines were applied to monthly means to interpolate daily values. The smoothing of rainfall and temperature data served to approximate the cumulative and delayed climatic conditions that shape malaria transmission [21] and lagged versions of both variables were generated to reflect the influence of prior rather than contemporaneous environmental conditions.

### Population data

Population estimates for the district of Moissala in 2023 were obtained from the *Direction des statistiques et du système d’information sanitaire (DSSIS)*, Ministry of Health, Chad. Because no age-stratified values were provided, the age distribution of Moissala was assumed to match that of Chad as a whole (19% of under 5 year old). To derive the 2018 population of Moissala, an annual growth rate for Chad of 0.0382, derived from United Nations population data [22], was applied in reverse to the 2023 estimate. This yielded an estimated Moissala population of approximately *P*_0_ = 103,000 in 2018, which served as the initial population size in the transmission model.

Similarly, to account for the effect of population growth on incidence, age-specific growth rates were derived using age-stratified population counts from the United Nations World Population Prospects[23]. The standard growth rate formula was used, 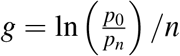, where *p*_0_ and *p*_*n*_ are the population sizes at the beginning and end of the period, and *n* is the number of days between 2018 and 2023. This resulted in daily growth rates of approximately 1.000079 for the under-five population and 1.000108 for those aged five years or older. These daily growth rates were used to adjust incidence estimates for population growth, as described in **Section** .

## Methods

### Malaria transmission model

#### Model structure

The malaria transmission model built on the framework proposed by Ukawuba and Shaman [9]. Several modifications were implemented to adapt the model to the epidemiological context of Moissala: (i) explicit incorporation of age structure, (ii) inclusion of the effects of SMC, and (iii) refinement of the climate–transmission relationships. The resulting model is a deterministic compartmental model stratified into two age groups: children under five years of age and individuals aged five years or older.

Within each age group, the population is distributed among five compartments: susceptible (S), exposed/incubating (E), infected and symptomatic but untreated (I^*U*^), infected, symptomatic, and treated (I^*T*^), and infected but asymptomatic (I^*A*^). Individuals age out of the under-five category and enter the older age group at a daily rate of 1*/*(5 *×* 365). A schematic of the compartmental structure is shown in **Figure 1**.

**Figure 1.**
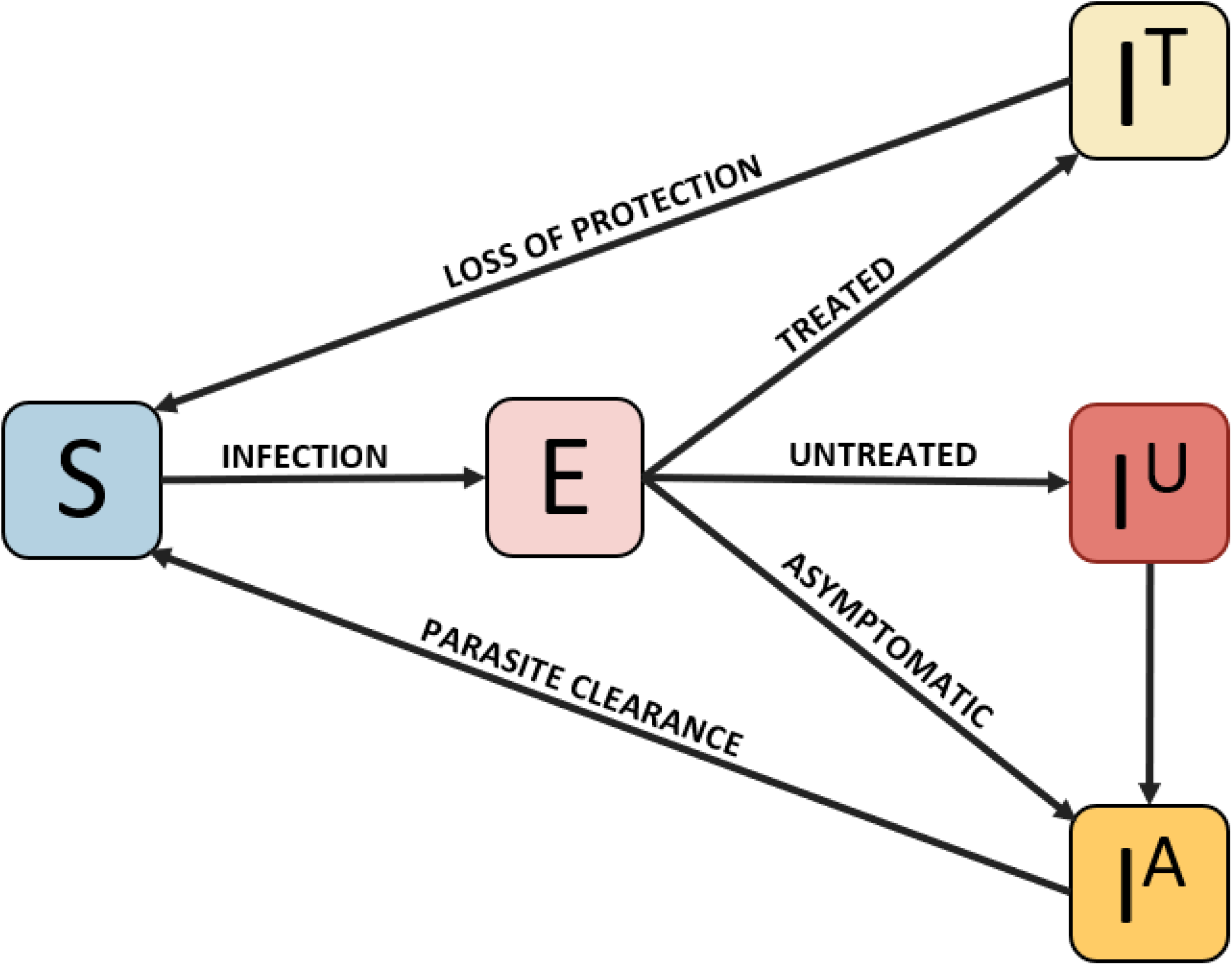
Diagram of the compartmental model. S: those susceptible to infection by malaria parasites, E: those who are infected by parasites (during incubation period), I^*U*^: those with symptomatic infection but remaining untreated, I^*T*^: those with symptomatic infection and treated, I^*A*^: those experiencing an asymptomatic infection. The model contains two age groups, those under 5 years of age and those 5 years or older. Seasonal malaria chemoprevention is administered to the under-five group and reduces the force of infection. Children transition from the under-five group to the older age group at rate 1*/*(5 *×* 365).

The model is defined formally by the following system of ordinary differential equations

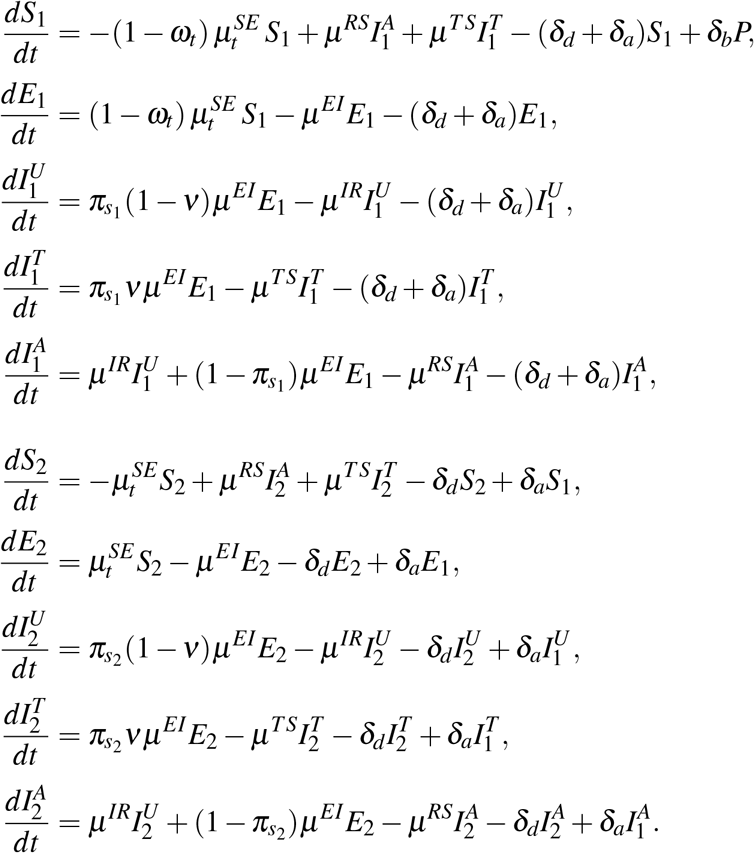

Where subscripts 1 and 2 denote the populations under and above five-year-old, respectively. Parameters *δ*_*b*_, *δ*_*d*_, and *δ*_*a*_ represent the birth, death, and aging rates. The term *ω*_*t*_ captures the time-varying protective effect of SMC. The probability of receiving effective treatment is denoted by *ν*, and 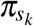 represents the probability that an infection becomes symptomatic (as opposed to asymptomatic) in age group *k*. Transition rates between compartments *X* and *Y* are represented by parameters of the form *µ*^*XY*^. All parameter definitions are provided in **Table 1** and **Table 2**.

**Table 1.** Fixed parameter descriptions.

| Name | Description | Unit | Value | Source |
| --- | --- | --- | --- | --- |
| **Immunity and Treatment Parameters** |  |  |  |  |
| $\pi_{s_1}$ | Proportion of all malaria infections that are symptomatic in those $< 5$ years old | Dimensionless | 0.86 | [26] |
| $\pi_{s_2}$ | Proportion of all malaria infections that are symptomatic in those $\geq 5$ years old | Dimensionless | 0.71 | Assumed to be equal to proportion in 5-15 age group reported in [26] |
| $v$ | Treatment rate | Dimensionless | 0.28 | [27] |
| $q_R$ | Infectivity of non-clinical cases relative to clinical cases | Dimensionless | 0.24 | [26, 28–30] |
| **Demographic Parameters** |  |  |  |  |
| $\delta_b$ | Birth rate | $\text{day}^{-1}\text{person}^{-1}$ | $47/(1000 \cdot 365)$ | **Appendix S2.2** |
| $\delta_d$ | Death rate | $\text{day}^{-1}\text{person}^{-1}$ | $47/(1000 \cdot 365)$ | **Appendix S2.2** |
| $\delta_a$ | Aging rate | $\text{day}^{-1}$ | $1/(5 \cdot 365)$ | - |
| $\kappa$ | Proportion of the population aged over five years | Dimensionless | 0.81 | [22] |
| **Transition Rates** |  |  |  |  |
| $\mu^{EI}$ | Parasite incubation rate | $\text{day}^{-1}$ | 1/10 | [31] |
| $\mu^{TS}$ | Loss of post-treatment prophylactic protection rate | $\text{day}^{-1}$ | 1/30 | [32–37] |
| $\mu^{IR}$ | Symptomatic-to-asymptomatic infection transition rate | $\text{day}^{-1}$ | 1/5 | [38–41] |
| $\mu^{RS}$ | Clearance rate of asymptomatic infection (return to susceptibility) | $\text{day}^{-1}$ | 1/195 | [42–46] |
| **Climate and Force of Infection Parameters** |  |  |  |  |
| $T_{\text{opt}}$ | The mean of the asymmetric Gaussian distribution used to model the EIR-temperature relationship | $^{\circ}\text{C}$ | 27.78 | **Appendix S2.1** |
| $R_{50\%}$ | Location parameter for the logistic function used to model the EIR-rainfall relationship | Dimensionless | 0 | - |
| $p_{MH}$ | Probability of transmission from mosquito to human | Dimensionless | 0.50 | [47, 48] |

**Table 2.** Estimated parameter descriptions. The rightmost column provides the median and 95% credible interval for the corresponding marginal posterior distribution.

| Name | Description | Unit | Prior | Median (95% CI) |
| --- | --- | --- | --- | --- |
| **SMC Force of Infection Reduction** |  |  |  |  |
| $\text{eff}_{\text{SMC}}$ | Relative effectiveness of seasonal malaria chemo-prevention on malaria transmission | Dimensionless | $\mathcal{U}(0, 1)$ | 0.69 (0.64, 0.76) |
| **Climate and force of infection Parameters** |  |  |  |  |
| $\sigma_{LT}$ | Standard deviation of the left side of the asymmetric Gaussian distribution used to model the EIR-temperature relationships. See **Appendix S2.1** for more details | $^{\circ}\text{C}$ | $\mathcal{N}(\mu = 3.20, \sigma = 1)$ | 3.90 (3.30, 4.61) |
| $\sigma_{RT}$ | Standard deviation of the right side of the asymmetric Gaussian distribution used to model the EIR-temperature relationships. See **Appendix S2.1** for more details | $^{\circ}\text{C}$ | $\mathcal{N}(\mu = 2.00, \sigma = 1)$ | 3.49 (3.34, 3.72) |
| $\tau$ | Rainfall sensitivity parameter in the EIR-rainfall relationship logistic function | Dimensionless | $\mathcal{U}(0, 10)$ | 1.71 (1.59, 1.80) |
| $j_1$ | Lag parameter for temperature | Dimensionless | $\mathcal{U}(0, 60)$ | 26 (23, 30) |
| $j_2$ | Lag parameter for rainfall | Dimensionless | $\mathcal{U}(0, 60)$ | 43 (40, 47) |
| $b$ | Saturation rate for the EIR-population infectivity relationship | Dimensionless | $\mathcal{U}(0, 200)$ | 2.88 (2.48, 3.00) |
| **Observation model parameters** |  |  |  |  |
| $s$ | Parameter accounting for uncertainty in population estimates | Dimensionless | $\mathcal{U}(0, 20)$ | 9.11 (8.18, 9.77) |
| $\phi$ | Dispersion parameter of negative binomial distribution | Dimensionless | $\mathcal{U}(0, 300)$ | 9.83 (9.16, 11.09) |

#### SMC effect

The overall effect of SMC at time *t*, denoted *ω*_*t*_, is modelled as a reduction in the force of infection experienced by children under five:

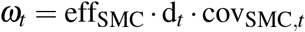

where eff_SMC_ is the intrinsic effectiveness of SMC, d_*t*_ is the time-varying drug efficacy factor, and cov_SMC,*t*_ is the estimated SMC coverage at time *t*. The parameter eff_SMC_ is inferred during model fitting and takes on values between 0 and 1, with values near 1 indicating strong protection among covered children and values near 0 indicating little or no protection.

The drug efficacy factor *d*_*t*_ incorporates the known decay of SMC protection over time, following the formulation of Milligan *et al*. [24]:

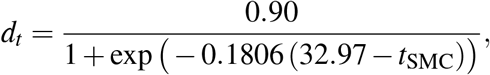

where *t*_SMC_ denotes the number of days since the start of the most recent SMC round. This function captures the rapid waning of drug-induced protection in the weeks following administration.

#### Force of infection

The force of infection at time *t*, denoted 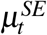 represents the daily probability that a susceptible individual becomes infected following exposure to infectious mosquito bites. Consistent with Ukawuba and Shaman’s climate-driven transmission framework [9], the force of infection is defined as: 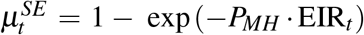, where *P*_*MH*_ is the probability that an infectious bite leads to human infection, and EIR_*t*_ is the entomological inoculation rate at time *t*, i.e. the expected number of infectious mosquito bites per person per day [25].

In the model, the EIR is governed by three components that capture key determinants of transmission: the level of infectiousness in the human population, local air temperature, and rainfall: 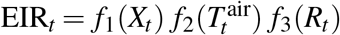.

#### Human infectiousness

The function *f*_1_(*X*_*t*_) = *X*_*t*_*/*(*b* + *X*_*t*_) relates the infection prevalence to transmission potential, where *X*_*t*_ = ∑_*k*_ (*I*_*k*,*t*_ + *q*_*R*_*R*_*k*,*t*_) */P* is the contribution to the EIR from the proportion of the population carrying either symptomatic or asymptomatic infections. Asymptomatic infections contribute less due to reduced infectiousness, governed by *q*_*R*_. This saturating functional form prevents unrealistically large EIR values when infection prevalence is high. The saturating constant *b* is inferred jointly with other model parameters.

#### Temperature–EIR relationship

Daily air temperature influences mosquito survival, parasite development, and the extrinsic incubation period. The original mechanistic relationships from [9] were replaced with a more flexible asymmetric Gaussian approximation to allow transmission to persist at the high temperatures observed in Moïssala, where daily values often exceed 32°C (see **Appendix S2.1**). In this formulation, air temperature enters through:

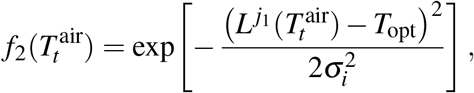

where 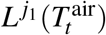 is the lagged air temperature value *j*_1_ days prior to time *t, T*_opt_ = 27.78^°^C is the optimal temperature for transmission, and *σ*_*i*_ takes the value *σ*_*LT*_ for temperatures below the optimum and *σ*_*RT*_ for temperatures above it. This asymmetric structure ensures a gradual decline in EIR at high temperatures, consistent with observed year-round malaria transmission in Moïssala. The values of *σ*_*LT*_ and *σ*_*RT*_ are inferred jointly with other model parameters.

#### Rainfall–EIR relationship

Rainfall influences the availability of larval habitats and mosquito abundance. Its effect is captured by a logistic function:

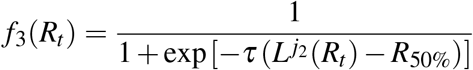

where 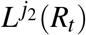 is the lagged rainfall value *j*_2_ days prior to time *t, τ* governs how sensitively EIR responds to rainfall changes, and *R*_50%_ represents the rainfall level associated with a 50% transmission probability. This formulation can capture the seasonal transitions in vector abundance characteristic of Moissala’s rainfall pattern. It should be noted that the function is monotonic, meaning that more rainfall always leads to increased transmission intensity.

### Inference

#### Observation model and model parameters

The reported (observed) number of malaria cases in children under 5 during epidemiological week *w* corresponds (in the transmission model) to the number of newly infected children that seek treatment during that week. This baseline weekly incidence is denoted by 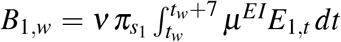, where *t*_*w*_ = 7(*w* − 1), where the subscript 1 denotes the under 5 age group, *ν* is the treatment seeking rate, *π*_*s*1_ the proportion of malaria infections that are symptomatic, and *µ*^*EI*^ is the parasite incubation rate. Because the model is implemented in discrete time with one-day steps, this integral is approximated by a sum over the seven daily time points.

To account for population growth, the baseline incidence was multiplied by an age-specific weekly growth rate *r*_1,*w*_. Let *g*_1_ = 1.000079 be the daily population growth rate for children under 5. The scaling factor for week *w, r*_1,*w*_, is obtained by raising the daily growth rate to the power of the number of days that has passed since week *w*. The final modelled reported cases for children under 5 in week *w* is then the baseline weekly incidence multiplied by the scaling factor, *C*_1,*w*_ = *r*_1,*w*_ *B*_1,*w*_.

A total of eleven parameters were estimated, namely the magnitude of SMC effectiveness, as well as parameters related to the climate-informed force of infection and the observation model (**Figure 2**).

**Figure 2.**
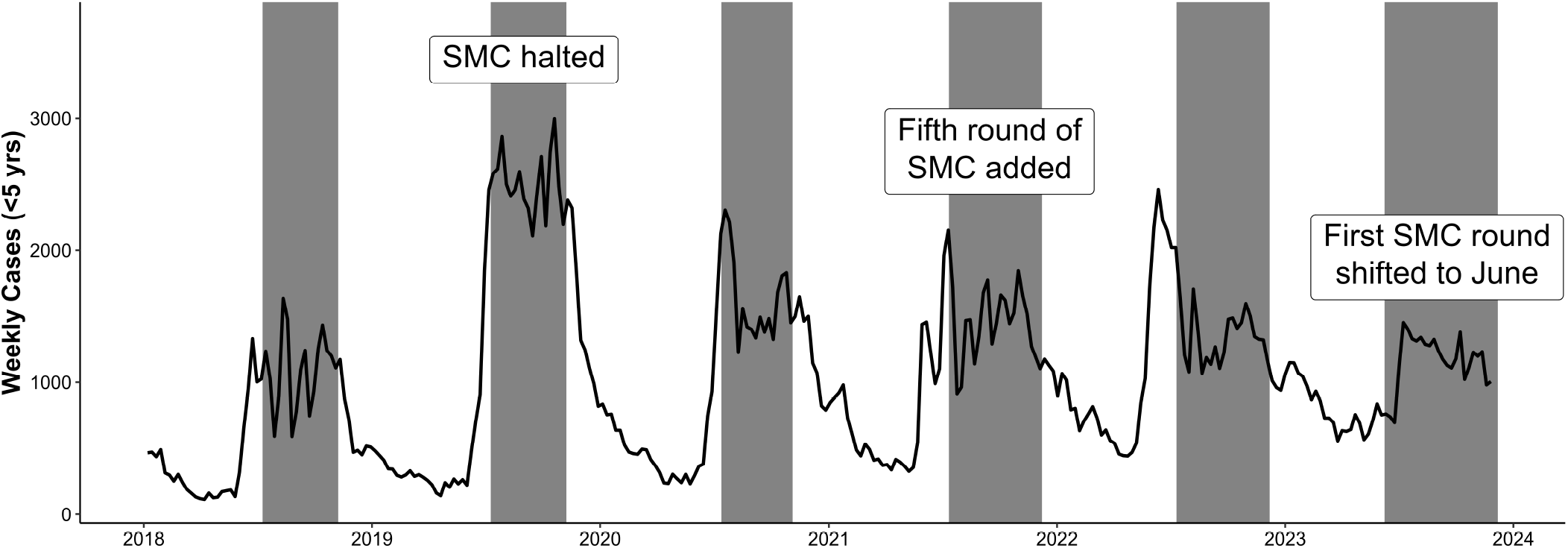
Graphical timeline of changing SMC strategies from 2018 to 2023. The black line represents the observed weekly malaria cases. Shaded recentagles are the time period when SMC was active.

To account for uncertainty in population size, an additional parameter *s* was introduced. This parameter scales the baseline population estimate linearly, such that 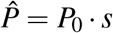, where *P*_0_ denotes the estimated population of Moissala in 2018. The introduction of *s* was necessary because the reported population size of 125000 in 2023, and the corresponding population size of 103000 used to initialize the model in 2018, is likely an underestimate of the true catchment population. This is due to the fact that the most recent population census was conducted in 2009, meaning current estimates are only projections, and to the likelihood that individuals residing outside Moissala seek treatment at health facilities within the district. As a result, the population effectively contributing to observed case counts may be larger than the provided population estimate. Including the parameter *s* therefore allowed the model to adjust the baseline population estimate to a level consistent with the scale of malaria transmission in Moissala.

A Bayesian inference framework was employed to estimate the model parameters. The prior distributions for each parameter were specified based on the available estimates derived from literature or previous work.

#### Adaptive MCMC procedure

The posterior distribution over the parameter set was estimated using an adaptive Metropolis–Hastings random walk algorithm, as implemented in the *mcstate* R package [49, 50]. The covaiance matrix of the proposal distribution is iteratively updated using the empirical covariance of the sampled parameters, improving sampling efficiency in the presence of strong parameter correlations.

To improve posterior exploration and ensure valid sampling, we employed a four-stage adaptive Metropolis–Hastings procedure (**Appendix S1.3)**. Stage 1 initialized chains from independent draws from uniform distributions, with lower and upper bounds chosen heuristically based on plausible ranges for each parameter. Each subsequent stage reused the final 3,000 samples from the preceding stage to compute the empirical covariance matrix, which informed the proposal distribution. The median of each parameter across these samples was also computed, and new starting values were drawn from a uniform distribution with bounds set to 0.8 and 1.2 times the median (reversed if the median was negative). Stage 2 increased the prior weight on the proposal covariance to stabilize adaptation. Stage 3 disabled adaptation entirely to inform the proposal for the final stage. Stage 4 continued sampling with this fixed proposal for 100000 iterations, discarding the first 20000 as burn-in to yield 80000 posterior draws.

### Implementation

The malaria transmission model and inference procedure are available as an open-source R package (https://github.com/SwissTPH/malclimsim). This package relies on *odin* [51], and *odin*.*dust* [52] for writing and simulating from the model and *mcstate* for parameter estimation [49, 53]. The ODEs were approximated in discrete time with a 1-day time-step. The package is accompanied by a tutorial vignette that guides a user through the process of preparing the climate and SMC input data, defining model inputs, inferring model parameters, and simulating from the model.

### Counterfactual simulations

The fitted model was used to simulate counterfactual scenarios to evaluate the effectiveness of SMC, the impact of having changed SMC strategies over time, and the optimal timing of SMC.

We first define some notation that will be used in the subsequent paragraphs: let *W* denote the set of weeks corresponding to the high transmission season from June to December, and let *Yr* denote the set of years included in the comparison. Let *N*_*Yr*_ = |*Y r*| denote the number of years in this set. The total number of cases under strategy *s* is defined as 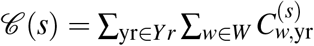, where 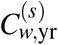 is the number of cases in week *w*, year yr, under strategy *s*.

Overall SMC effectiveness was assessed by comparing the implemented SMC strategy to a counterfactual strategy in which SMC was not implemented in any year. Let *s* ∈ {implemented, no SMC}, and let *Yr* = {2018, 2020, 2021, 2022, 2023}, excluding 2019 when SMC was not implemented. Effectiveness is then defined as 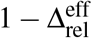, where 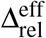 denotes the ratio of total cases under the implemented strategy to those under the no-SMC counterfactual and is given by 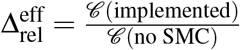. Yearly cases averted due to SMC is the difference in cases under the no SMC strategy and the implemented strategy, defined as 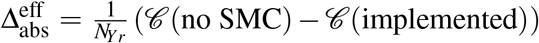.

The fitted model was then used to assess the impact of three specific changes to the SMC strategy: (1) halting SMC in 2019, (2) adding a fifth round in 2021, and (3) shifting the first round to June. A graphical timeline of these strategies is shown in **Figure 2**. For each change, two simulations were conducted: an observed scenario reflecting the implemented strategy and coverage, and a counterfactual scenario identical in all respects except that the SMC strategy in the affected year(s) reverted to the previously used approach. For instance, to assess the effect of halting SMC in 2019, the counterfactual simulation retained the four-round schedule beginning in July, as implemented before 2019. In this setting, *s* ∈ {implemented, counterfactual}, and *Yr* denotes the set of years affected by the strategy change. The absolute impact of a strategy change was defined as 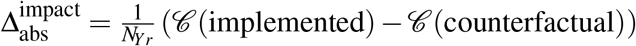 and the relative impact as 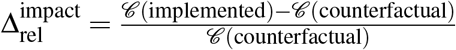.

The model was also used to evaluate which SMC strategy may be optimal across all years from 2018 to 2023. Eight strategies were compared, varying by the number of rounds (four or five) and the start date of the first round (June 1st, June 15th, July 1st, or July 15th). Coverage was fixed at 85%, corresponding to the average coverage across all months in which SMC was implemented.

Using the same notation, let *s* ∈ {strategy, baseline}, where “strategy” refers to the candidate SMC strategy and “baseline” to the historical strategy of four rounds of SMC starting on July 15th. Let *Yr* = {2018,…, 2023}. The absolute difference in cases was defined as 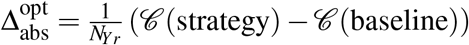.

## Results

### Model calibration and estimation of SMC effectiveness

The malaria transmission model was calibrated using weekly data on the number of malaria cases in children under 5 years old from January 2018 to December 2023, as shown in **Figure 3**. The model captures the overall trends in reported cases, with all observations comprised in the 95% prediction intervals. Some discrepancies were observed in specific years: the peak in 2019 is underestimated, and the number of cases in 2023 is overestimated.

**Figure 3.**
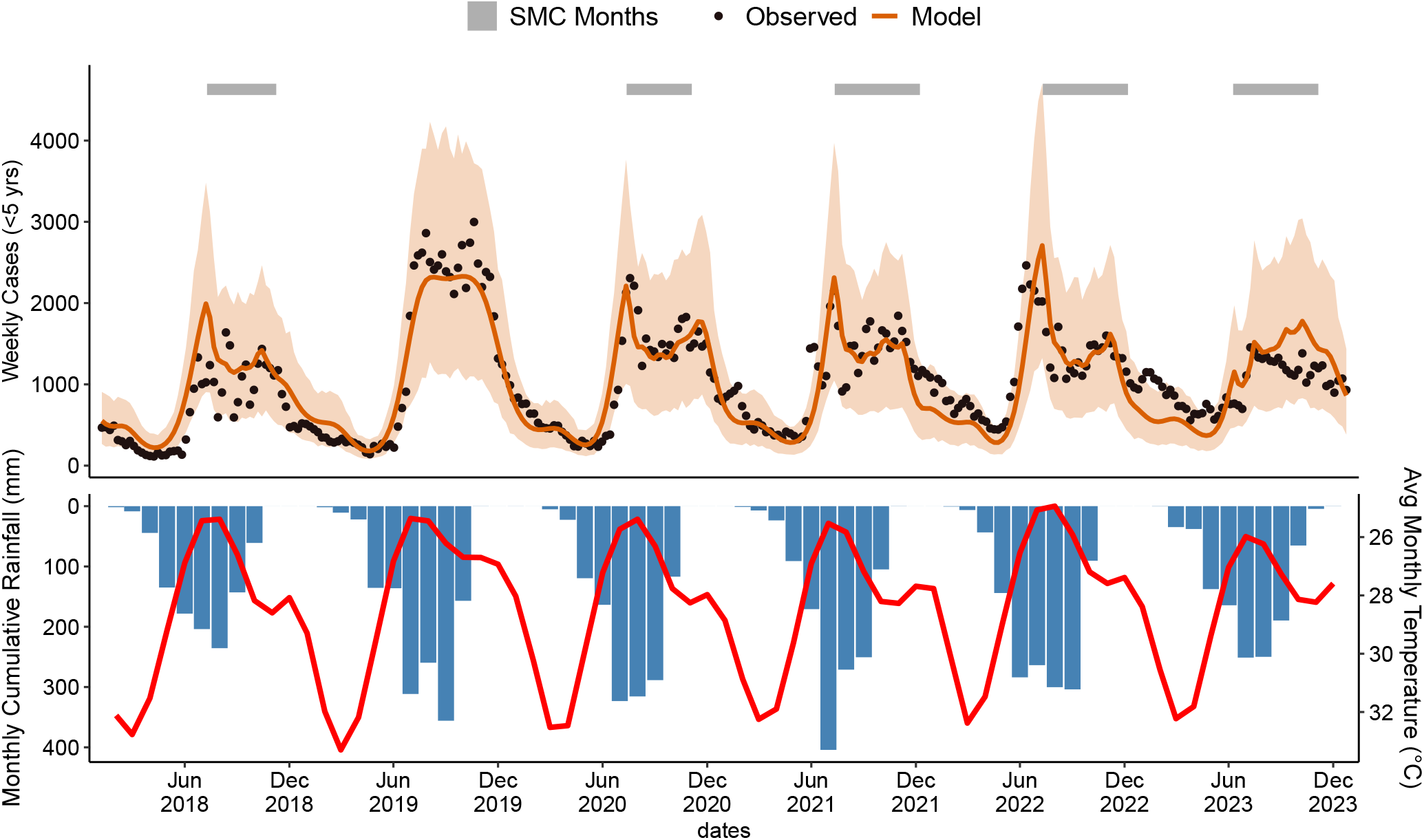
Comparison of observed and modeled weekly malaria cases in children under 5 years old in Moissala, Chad from 2018 to 2023. Black dots show observed cases. The solid orange line shows model predictions under the maximum a posteriori (MAP) parameter set, and the orange band denotes the 95% prediction interval based on 1000 posterior draws. Prediction intervals reflect uncertainty about future observations and are therefore wider than its corresponding credible interval, which reflect uncertainty about parameters. Gray bands indicate months during which SMC was implemented.

The effectiveness of SMC from 2018 to 2023 was assessed using several metrics. The first is the estimated value of the eff_SMC_ parameter of the malaria transmission model, representing the reduction in force of infection in children under 5 years old due to SMC. The estimated value was 68% (95% CI: 60%, 76%) (**Table 3**). The calibrated model was used to simulate weekly cases in children under 5 years old from 2018 to 2023, excluding 2019 when SMC was not implemented, under two strategies: SMC each year with coverages equal to the observed coverages and a strategy of no SMC during the entire time period (**Figure 4**). An estimated 14400 (95% CI: 12000, 16800) cases in children under 5 years old were averted each year during the high transmission season due to SMC, corresponding to an effectiveness of 26% (95% CI: 21%, 31%) (**Table 3**).

**Table 3.**
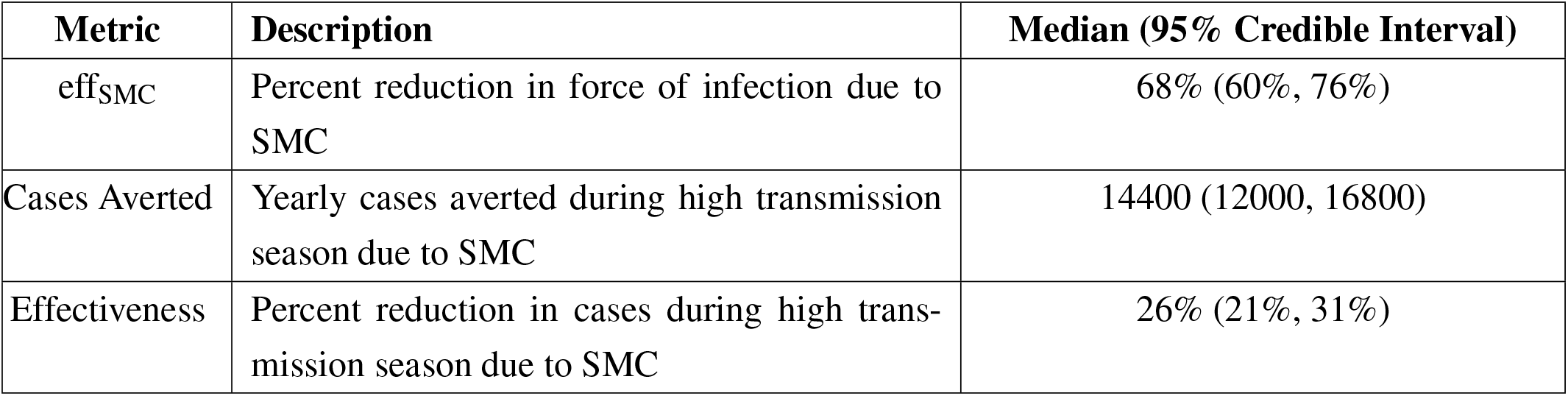
Estimated effectiveness of SMC in children under 5 years old in Moissala, Chad during the high transmission season (June to December). The table gives the posterior median and 95% credible interval for the transmission model parameter that governs reduction in force of infection eff_SMC_, the estimated number of malaria cases averted per year due to SMC, and the corresponding percent reduction in cases relative to a counterfactual where SMC was not implemented. Further details can be found in **Section**. Medians and credible intervals are derived from simulations using 1000 parameter sets drawn from the posterior distribution.

| Metric | Description | Median (95% Credible Interval) |
| --- | --- | --- |
| $\text{eff}_{\text{SMC}}$ | Percent reduction in force of infection due to SMC | 68% (60%, 76%) |
| Cases Averted | Yearly cases averted during high transmission season due to SMC | 14400 (12000, 16800) |
| Effectiveness | Percent reduction in cases during high transmission season due to SMC | 26% (21%, 31%) |

**Figure 4.**
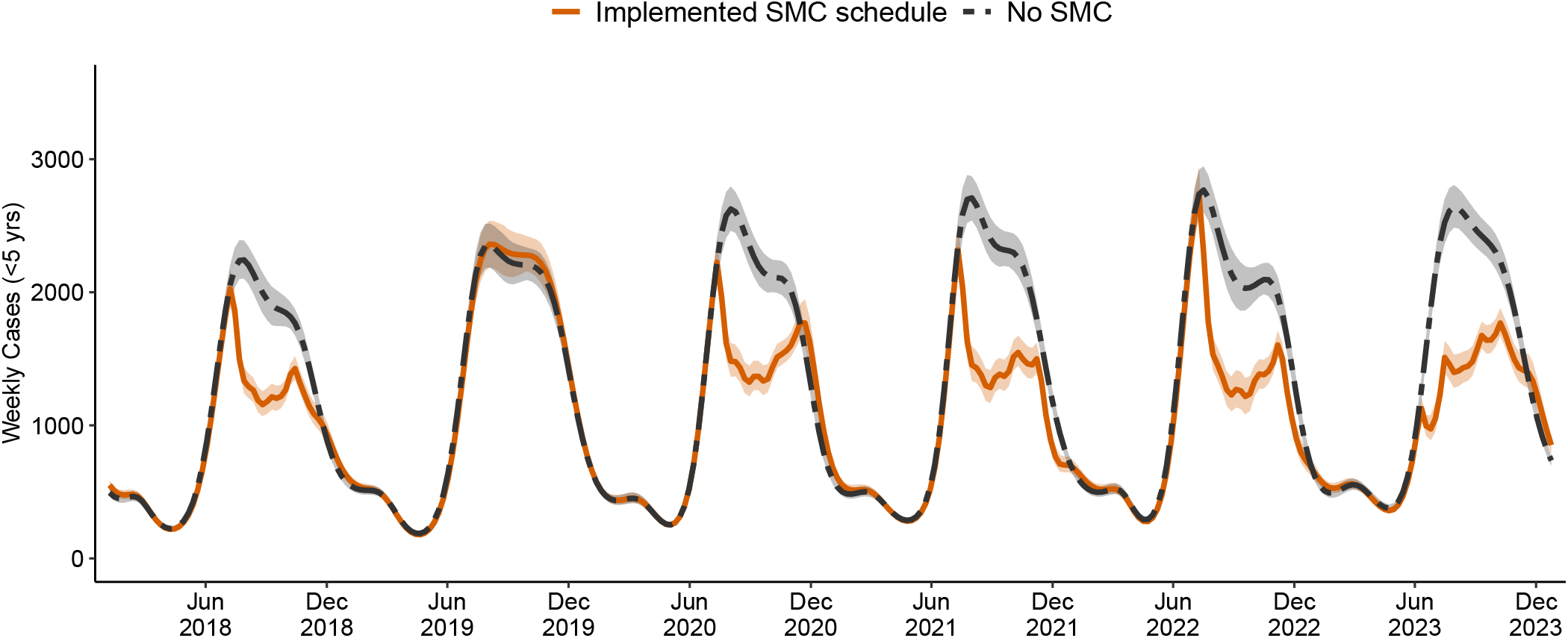
Modelled weekly malaria cases in children under five in Moissala, with and without seasonal malaria chemoprevention (SMC), January 2018 to December 2023. The solid orange line shows model simulations under the historically implemented SMC schedule; the dashed gray line shows the counterfactual scenario with no SMC. Shaded bands around each line indicate the 95% credible interval derived from 1000 posterior model draws, reflecting uncertainty in the modelled mean number of cases per week.

### Impact of SMC strategy changes

To assess the impact of changes to the SMC schedule on malaria burden in Moissala, simulated weekly malaria cases in children under 5 years old under the implemented strategy were compared to a counter-factual scenario in which the previous SMC strategy was retained during the years affected by a change (**Figure 5**). For example, to evaluate the effect of discontinuing SMC in 2019, the number of cases in 2019 was simulated both under the observed strategy of no SMC and under the historical strategy of four rounds of SMC beginning in July. Comparing the two simulations provides the estimated impact of the change.

**Figure 5.**
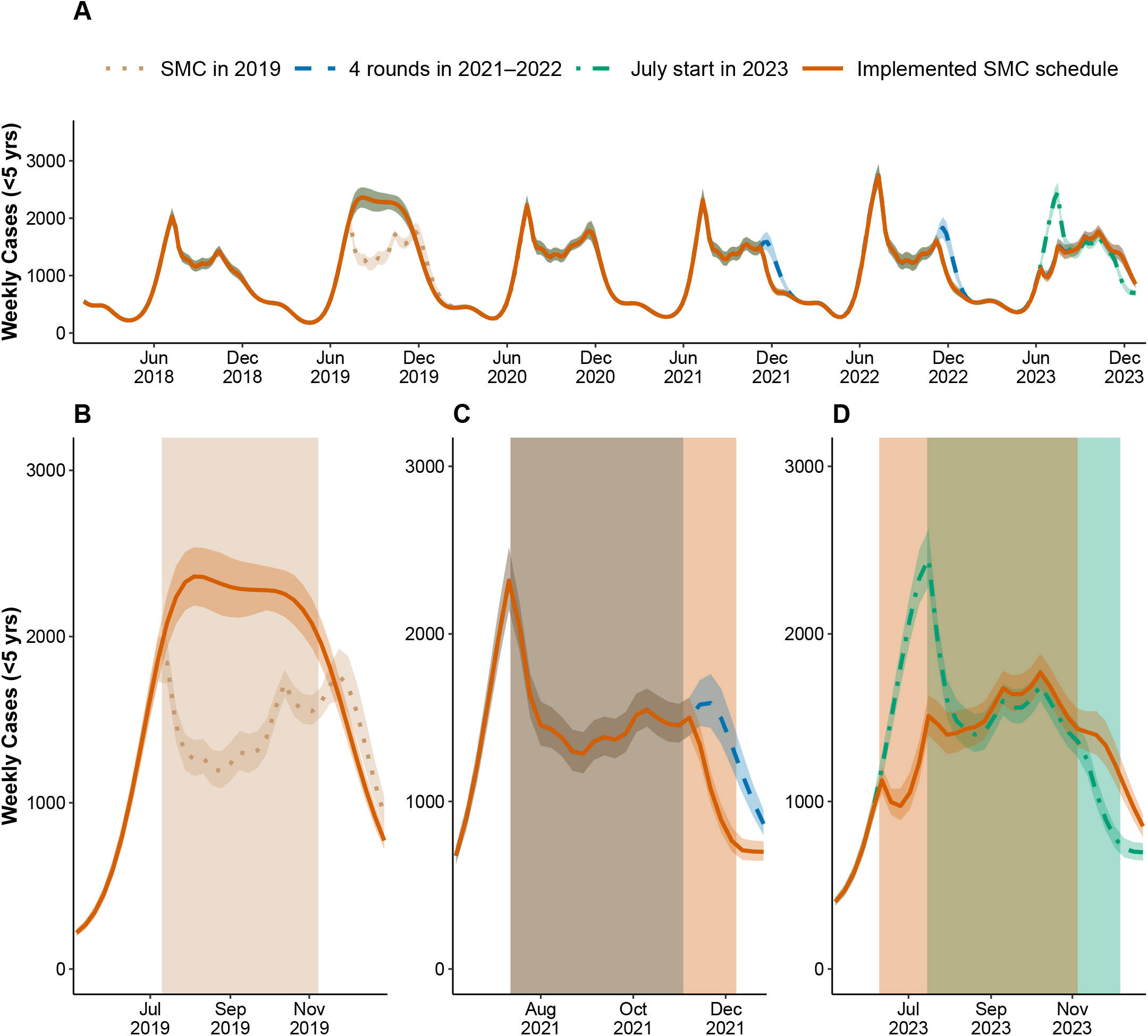
Modelled impact of SMC strategies on malaria burden in children under 5 years old from 2018 to 2023. **A:** Comparison of the implemented SMC strategy to counterfactuals representing what would have happened had the most recent strategy been continued. **B-D:** Each plot corresponds to a year a change in SMC strategy occurred. **B:** SMC was discontinued. **C:** A fifth round of SMC was added in November. **D**: The first round of SMC was shifted to June. Shaded bands around each line indicate the 95% credible interval derived from 1000 posterior model draws, reflecting uncertainty in the modelled mean number of cases per week. The shaded rectangles are the months where SMC was active for a given strategy.

Discontinuing SMC in 2019 resulted in an estimated increase of 13600 cases (95% CI: 11200 to 15800) in children under 5 years old, corresponding to a 31% increase (95% CI: 25% to 38%) (**Table 4**). Adding a fifth round of SMC at the end of the transmission season in 2021 and 2022 reduced cases in children under 5 years old by 3,000 per year (95% CI: 2400 to 3600), or 7% (95% CI: 5% to 8%). Advancing the start of SMC to June in 2023 is estimated to have reduced cases in children under 5 years old by 2100 (95% CI: 1600 to 2800), or 5% (95% CI: 4% to 6%), compared to if the five rounds had started in July.

**Table 4.** Estimated impact of changes to the SMC schedule on weekly malaria cases in children under 5 years old. The counterfactual is the SMC implementation that was done until the change occurred. Absolute reports the median and 95% credible interval for yearly case differences (implemented strategy minus counterfactual). Relative reports the percent change in cases of the implemented strategy compared to the counterfactual strategy. The relative change is calculated as difference in yearly cases between implemented strategy and counterfactual, divided by the yearly cases for the counterfactual strategy, multiplied by 100. The number of cases is reported as yearly cases. Further details can be found in **Section** .

|  |  | 2019 | 2021–2022 | 2023 |
| --- | --- | --- | --- | --- |
| **Implemented strategy** |  | No SMC | 5 Rounds July | 5 Rounds June |
|  | Cases | 56600 (54100, 59000) | 41800 (39900, 43800) | 40900 (38800, 43300) |
| **Counterfactual** |  | 4 Rounds July | 4 Rounds July | 5 Rounds July |
|  | Cases | 4300 (41100, 44800) | 44800 (43000, 46600) | 43100 (41200, 45300) |
| **Change** | Absolute | 13600 (11200, 15800) | -3000 (-3600, -2400) | -2100 (-2800, -1600) |
|  | Relative (%) | 31% (25%, 38%) | -7% (-8%, -5%) | -5% (-6%, -4%) |

### Optimal SMC timing

To identify the most effective SMC strategy, a range of scenarios was simulated, varying in the number of SMC rounds, the starting month, and the start date within the month. Each strategy was compared to a baseline scenario of four rounds of SMC starting on July 15th. Coverage was held at 85% across simulations (**Figure 6A**), the estimate average SMC coverage from 2018 to 2023.

**Figure 6.**
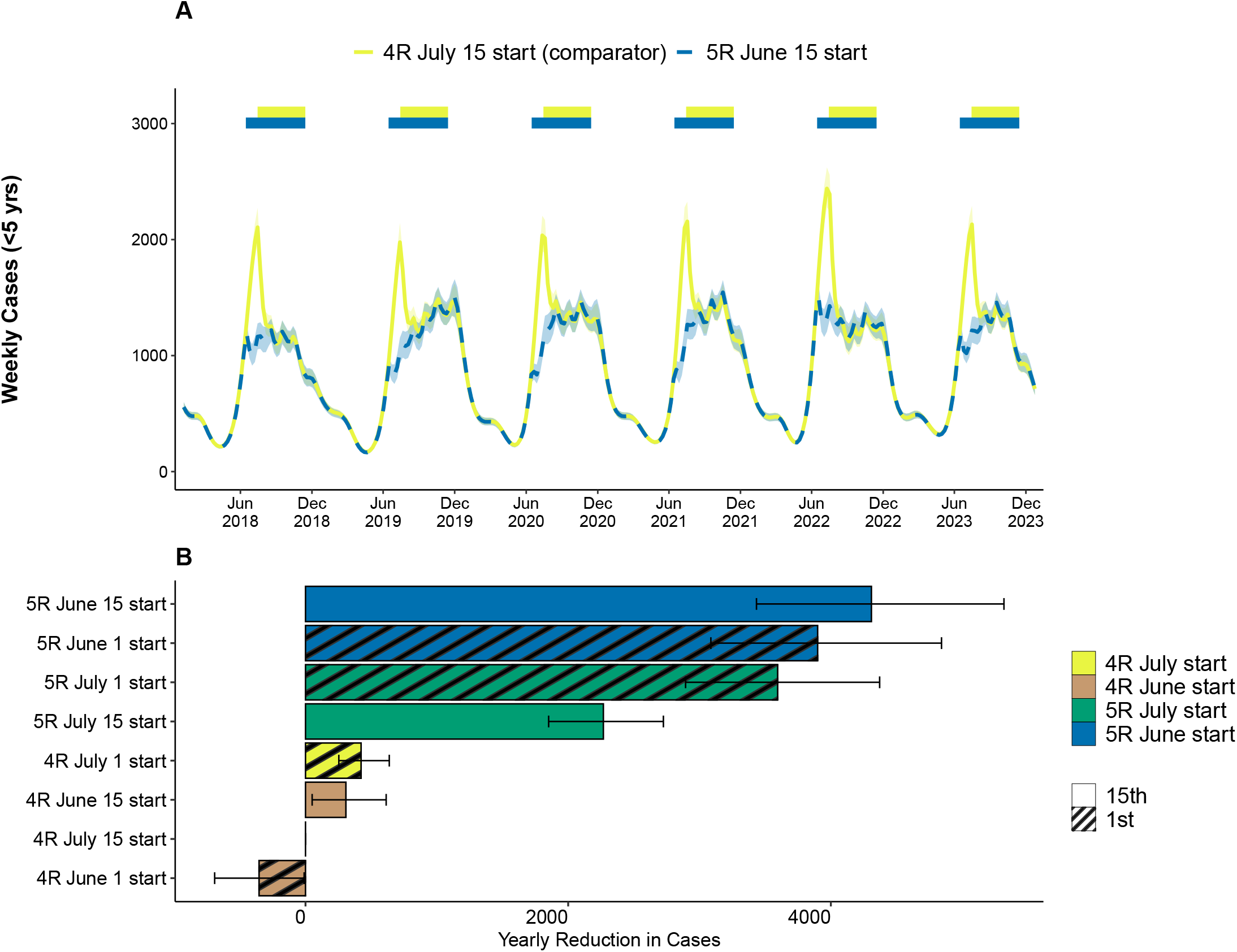
Determining the optimal SMC strategy. Each strategy was compared to the historical strategy of four rounds of SMC with the first round of SMC starting in July. **A:** Comparison of the historical strategy with the strategy that led to the greatest reduction in cases in children under 5 years old. Shaded bands around each line indicate the 95% credible interval derived from 1000 posterior model draws, reflecting uncertainty in the modelled mean number of cases per week. **B:** Comparison of each strategy to the historical strategy. The bar length is the median and the error bar gives the 95% credible interval using parameter sets from 1000 draws of the posterior distribution.

The greatest reduction in malaria cases was observed under the strategy of five rounds of SMC beginning on June 15th, with 4300 (95% CI: 3400 to 5200) fewer cases in children under 5 years old per year relative to the baseline of four rounds of SMC beginning on July 15th (**Figure 6B**). The next most effective strategies were five rounds of SMC beginning on June 1st, July 1st, and July 15th. Among the four-round strategies, starting on July 1st reduced cases the most, with 420 (95% CI: 250 to 640) cases being averted in children under 5 years old per year compared to starting on July 15th. A complete summary of all strategies evaluated is provided in **Table S1.4**.

## Discussion

Using a climate-driven model of malaria transmission calibrated to routine case data, we estimate that the implementation of SMC in Moissala led to a 26% reduction in malaria cases in children under 5 years old during the high transmission season from 2018 to 2023 and that children who received the intervention were on average 68% less likely to be infected. We also evaluated changes to the SMC strategy over time. Specifically, halting SMC in 2019 resulted in a 31% increase in malaria cases in children under 5. On the contrary, adding a fifth round and advancing the start date, contributed to reductions in malaria burden by 7% and 5% respectively. Our results suggest that the optimal strategy, accounting for inter-annual climate variation, is five rounds of SMC beginning on June 15th. This recommendation aligns with the current strategy in use.

The conclusions of this work have been used by stakeholders to guide SMC strategy in terms of the number of rounds and timing to optimize the SMC implementation in Moissala. This analysis also demonstrates how tailoring intervention strategies to the geographical context, leveraging the available data, can help further reduce the malaria burden. Additionally, it confirms the relevance of SMC in areas with a long transmission season, in line with the evolution of the WHO SMC guidelines. This evidence can therefore be used to support the roll-out of SMC with five rounds in the southern part of Chad where the transmission season is longer.

Our estimate of SMC effectiveness under routine implementation is in line with the published literature. It is slightly higher than the estimate reported by Richardson [54], which found a 19% reduction in Chad over the 2013 to 2018 period. Estimates from a paper published as part of the ACCESS-SMC project, which analyzed reductions in confirmed outpatient cases during the high transmission season across seven countries, reported a 44% reduction in malaria cases in children under 5 in Chad across 2015 and 2016 [4]. Other country-specific estimates ranged from 26% in Nigeria to 55% in The Gambia. While the effectiveness calculation used in [4] is broadly similar to ours, they focused exclusively on cases occurring between August and November (the peak transmission months in those particular contexts). Restricting our analysis to that same period yields an estimated reduction of 32%, which is more closely aligned with their results.

Our estimate is substantially lower than those reported in randomized controlled trials (RCTs). A recent meta-analysis of twelve RCTs estimated an average reduction in malaria incidence of 73%, with individual trial estimates ranging from 38% to 86% [55–66]. This discrepancy reflects the known gap between efficacy under trial conditions and effectiveness under real-world implementation, where factors such as lower adherence, coverage gaps, and operational inefficiencies may reduce impact [67]. These findings underscore the importance of measuring effectiveness under real world conditions. Our approach provides a framework for doing so in a robust and timely manner.

Our model incorporates real-time climate data, as this driver of malaria transmission could otherwise be an important confounder when estimating the effectiveness of SMC. For example, in assessing the impact of halting SMC in 2019, it is observed that rainfall increased compared to previous years. It follows that part of the rise in cases is indeed likely due to the discontinuation of SMC, but another part is likely due to increased rainfall. Not accounting for rainfall in this case could result in an overestimation of the intervention’s impact. Climate also plays a central role in determining the optimal strategy. The best strategy is influenced not only by the total amount of rainfall, but also by the timing of the onset of the rainy season. In future analyses, this model could also be used to assess the impact of climate change on both malaria transmission and SMC impact in the area.

The model simulation and inference procedure is distributed as an open-source R package on GitHub (https://github.com/SwissTPH/malclimsim), making it accessible to a broad range of users. In addition, there are accompanying tutorials that guide users through the package’s functionalities. This facilitates the application of the methodology to other geographies, allowing users to estimate the impact of SMC and determine the optimal strategy in a specific context. Given the increased flexibility of the WHO’s SMC recommendations and growing interest from PNLPs in tailoring SMC strategies to their specific climatic conditions, we believe this represents an important contribution of our work.

Our model was well-adapted for our purpose, but several limitations should be noted. First, we simplified immunity by only differentiating between children under five and those aged five or older in terms of risk of developing symptomatic infection; other important immunity dynamics, such as immunity resulting from previous parasite exposure, or life-history of the disease such as superinfection or recurring bouts of fever, are not represented.

Interventions other than SMC and routine case management were not modelled directly. This relied on the assumption that there was little variability in the other prevention tools as the region did not yet benefit from indoor residual spraying or malaria vaccines at the time of study period. However, regular insecticide-treated net (ITN) distributions were conducted every 3 years. If ITN coverage and use varied substantially across years, this could bias the estimated impact of SMC. Future work could involve the exploration of ITN usage in Moissala as well as the best method to incorporate it into our framework.

Climatic drivers were limited to rainfall and temperature, which were included mechanistically based on field-informed relationships. Other environmental factors, such as humidity, vegetation, or extreme weather events like flooding, could also influence malaria transmission dynamics but were not explicitly modelled [68–71].

As in many applied modelling efforts, the model relies on routine health facility data. Due to its granularity in space and time, routine data is a powerful source of information to inform public health decision, allowing decision-makers to respond more quickly and more precisely to changes in malaria epidemiology [72–74]. However, routine data is also subject to reporting delays, underreporting, and other systemic challenges [15, 17, 75–77]. Although data quality constraints are common in programmatic contexts, they can bias the modelled impact, particularly if these challenges change over time. In our study, the decision to suspend SMC in 2019, which was not triggered by trends in malaria transmission, constitutes a “natural experiment” [78], and strongly reduces these biases. Nonetheless, we rely on a single time series to evaluate different counterfactual strategies and we observe only a single year without SMC implementation. Any unobserved change in surveillance, health-seeking, or transmission in that year could bias estimates of impact. In future work, incorporating control districts that are similarly affected by such changes could help reduce concerns about bias.

Uncertainty in SMC coverage data was not propagated through to the impact estimates. Because the results focused on relative comparisons using coverage levels that did not vary substantially over time, this omission is unlikely to affect qualitative conclusions. However, future work aiming to assess the absolute impact of varying coverage levels would benefit from incorporating uncertainty in coverage estimates explicitly.

Finally, we limited our analysis to uncomplicated cases in children under five, both for estimating impact and determining the optimal strategy. While older age groups are not targeted for SMC in Moissala, excluding them may lead to underestimation of overall impact: there is some evidence that suggests the presence of indirect effects following SMC implementation, potentially due to overall reduced transmission [79]. As a result, this analysis did not explore the impact of SMC age extensions, a question that can be better addressed with individual-based models [80].

## Conclusion

In conclusion, this work contributes to the evidence base on SMC effectiveness, demonstrates how climate-driven modelling can support programmatic decisions, and provides a framework for evaluating and optimizing SMC strategies. The methodology is implemented in a user-friendly, open-source R package, which facilitates future applications in other geographies. By enabling faster, locally informed decision-making, this work supports efforts to improve tailoring of SMC implementation to suit local malaria epidemiology, ultimately reducing the malaria burden.

## Data Availability

The malaria transmission model is available as an open source R package at https://github.com/SwissTPH/malclimsim. Code used to apply the model and reproduce the results of the manuscript can be found at https://github.com/epicentre-msf/smc-modelling-moissala. Climate, SMC coverage, and malaria incidence data will be made available in a Zenodo repository.

## Acknowledgments

Calculations were performed at the sciCORE (http://scicore.unibas.ch/) scientific computing core facility at the University of Basel.

This work was supported, in whole or in part, by the Gates Foundation [INV-068864] (EP, NP, IU, CC). The conclusions and opinions expressed in this work are those of the author(s) alone and shall not be attributed to the Foundation. Under the grant conditions of the Foundation, a Creative Commons Attribution 4.0 License has already been assigned to the Author Accepted Manuscript version that might arise from this submission. Please note works submitted as a preprint have not undergone a peer review process.

## Data and code availability

The model is available as an open source R package at https://github.com/SwissTPH/malclimsim. Code used to apply the model and reproduce the results of the manuscript can be found at https://github.com/epicentre-msf/smc-modelling-moissala. Climate, SMC coverage, and malaria incidence data will be made available in a Zenodo repository.

