## Supplementary material for "Modelling malaria routine surveillance data to inform seasonal malaria chemoprevention strategy in Moissala, Southern Chad"

[1]Swiss Tropical and Public Health Institute, Basel, Switzerland
[2]University of Basel, Basel, Switzerland
[3]Epicentre, Paris, France
[4]Médecins Sans Frontières, Paris, France
[5]Programme National de Lutte contre le Paludisme, Chad
[6]Clinton Health Access Initiative, Boston, USA
[*]equal contribution

### S1 Supplemental figures and tables

#### S1.1 MCMC diagnostics

**Figure S1:** MCMC trace plots and posterior distributions. **A:** Trace plots resulting from the fourth stage of the inference procedure. A total of four chains were initiated and ran for 80,0000 steps each. **B:** Marginal posterior distributions for each of the estimated parameters. Shaded region is the prior distribution.

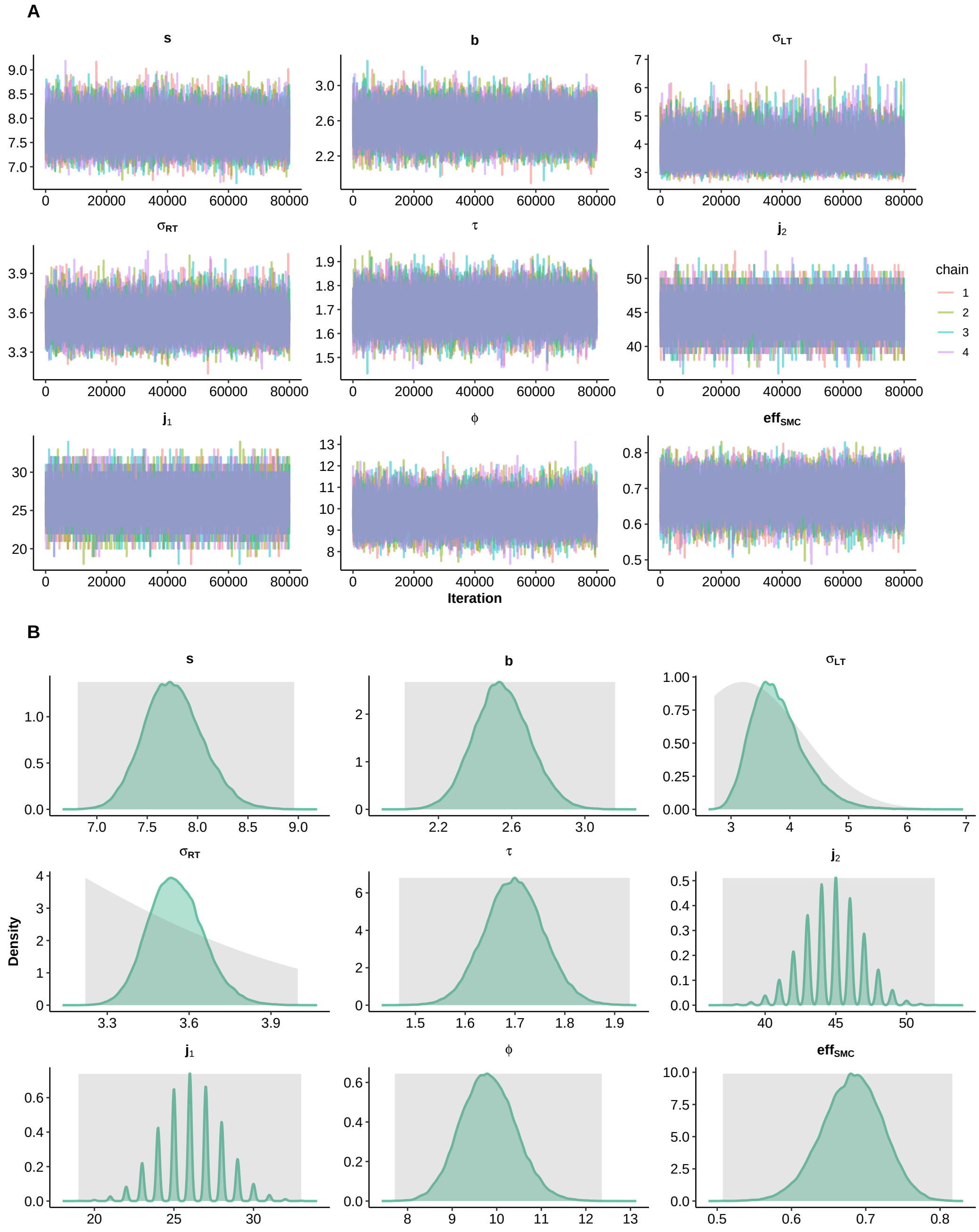

#### S1.2 SMC coverage data

**Figure S2:** Seasonal malaria chemoprevention coverage by year and round estimated from household coverage surveys conducted by MSF. The length of the bar is the mean and error bar gives the 95% confidence interval.

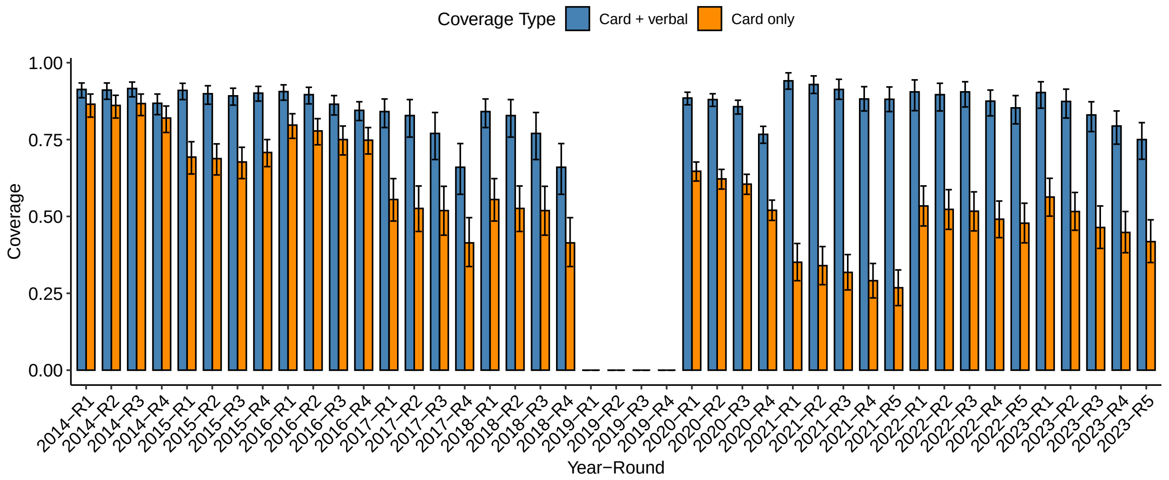

#### S1.3 Adaptive Metropolis-Hastings configuration

**Table S1:** Configuration of adaptive Metropolis–Hastings parameters across MCMC stages. Parameters marked **NA** are not relevant once adaptation is fully disabled via `adapt_end = 0`. A value of ∞ for `adapt_end` and `forget_end` indicate that covariance matrix adaptation and the forgetting of earlier parameter sets are always active. Further details of each algorithm parameter can be found in the documentation of *mcstate* [1].

| **Parameter** | **Stage 1** | **Stage 2** | **Stage 3** | **Stage 4** |
|---|---|---|---|---|
| *Scaling of proposal distribution* | | | | |
| Initial VCV weight | 1 | 100 | 500 | 500 |
| Initial scaling | 2 | 2 | 1 | 1 |
| Min scaling | 0.1 | 0.1 | 0 | 0 |
| Acceptance target | 0.234 | 0.234 | NA | NA |
| *Adaptation and forgetting of empirical covariance* | | | | |
| Adapt end | ∞ | ∞ | 0 | 0 |
| Forget rate | 0.6 | 0.4 | NA | NA |
| Forget end | ∞ | ∞ | NA | NA |
| Pre-diminish | ∞ | 20000 | NA | NA |
| *MCMC control* | | | | |
| Steps | 20000 | 60000 | 20000 | 100000 |
| Burn-in | 0 | 0 | 0 | 20000 |
| Chains | 4 | 4 | 4 | 4 |

#### S1.4 Additional optimal strategy figures and tables

**Figure S3:** Visual comparison of different SMC strategies from 2018 to 2023. **A:** Comparison between baseline strategy and strategies starting on the 15th of the month. **B:** Comparison between baseline strategy and strategies starting on the 1st of the month. The yellow, solid line is the baseline SMC strategy of four rounds of SMC with the first round on July 15th. A constant SMC coverage of 85% was used for each round. The difference between a strategy's trajectory and the baseline trajectory represents the excess cases or cases averted by that strategy.

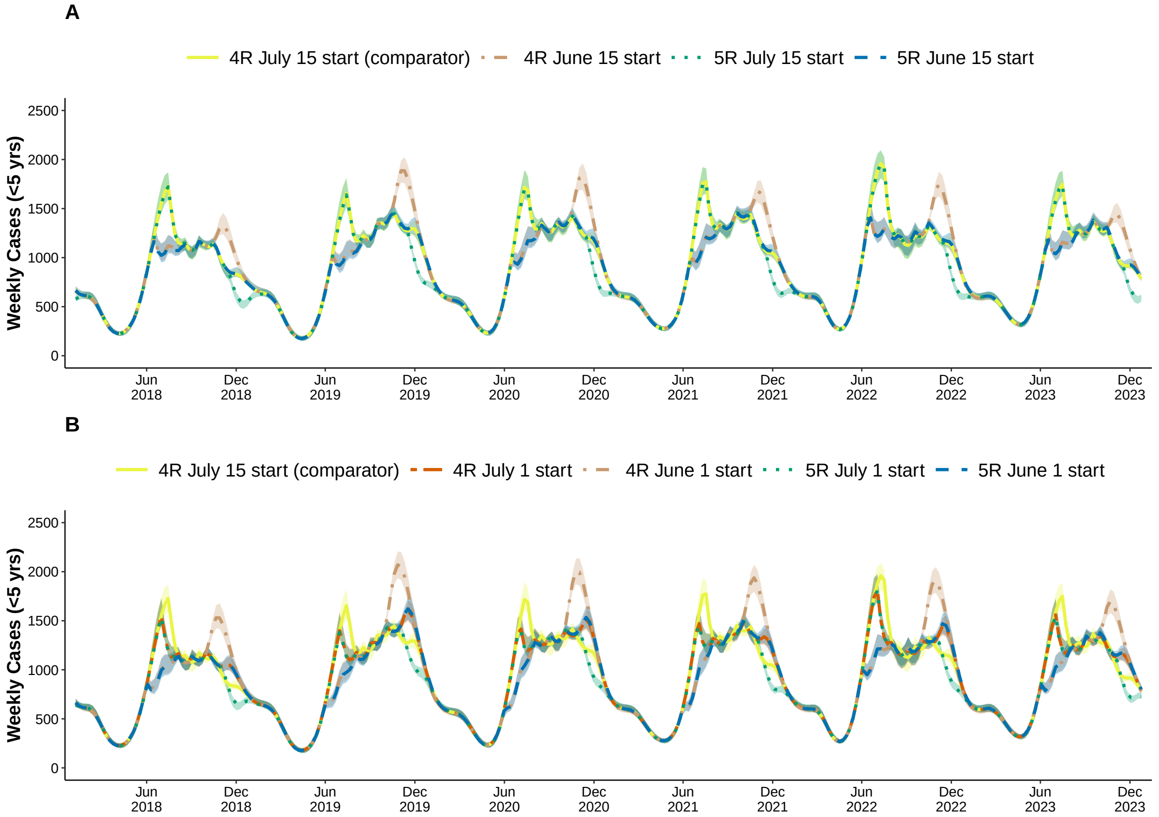

| **Number of Rounds** | **Start Date** | **Absolute Change in Cases** (per year) | **Relative Change %** |
|---|---|---|---|
| 5 Rounds | June 15th | -4300 (-5300, -3400) | -9% (-11%, -7%) |
| 5 Rounds | June 1st | -3900 (-3100, -4800) | -8% (-10%, -7%) |
| 5 Rounds | July 1st | -3600 (-4400, -2900) | -7% (-9%, -6%) |
| 5 Rounds | July 15th | -2300 (-2700, -1900) | -5% (-6%, -4%) |
| 4 Rounds | July 1st | -420 (-640, -250) | -1% (-1%, 1%) |
| 4 Rounds | June 15th | -300 (-610, -50) | -1% (-1%, 0%) |
| 4 Rounds | July 15th | 0 (0,0) | 0% (0%, 0%) |
| 4 Rounds | June 1st | 350 (10, 690) | 0% (0%, 0%) |

**Table S2:** Optimal SMC timing. The median and credible interval are estimated using parameter sets from 1000 draws of the posterior distribution. The absolute change in cases is a result of taking the difference between the total number of cases from 2018 to 2023 in strategy being compared to the comparator strategy (4 rounds of SMC with the first round in July) and then dividing by the total number of years in the analysis. The relative change is calculated as the absolute change in cases, divided by the yearly cases for the comparator strategy, multiplied by 100.

### S2 Supplemental text

#### S2.1 Simplification of climate equations

The mechanistic transmission model proposed for the analysis closely followed Ukawuba and Shaman [2], who calibrated temperature–EIR relationships for Rwanda. Unlike Rwanda's relatively mild, year-round climate, the Moïssala district in southern Chad experiences extreme summer temperatures (> 32 degrees Celsius) and a pronounced dry season. Applying the unmodified Ukawuba and Shaman temperature functions to Moïssala produced implausibly low EIRs for much of the year, because each entomological rate (survival probability, development time, extrinsic incubation period) is governed by a curve that drops to zero (or diverges) past a threshold temperature. Consequently, predicted EIR falls to zero above around 31.8 degrees Celsius, despite malaria cases being reported year-round (**Figures S4C, S4D**).

To retain the mechanistic intepretation but allow transmission beyond 32 °C, we approximated the Ukawuba and Shaman EIR–temperature curve with a two-sided (asymmetric) Gaussian. The curve is centered at the optimal temperature, $T_{\text{opt}} = 27.78$ and uses two separate shape parameters, $\sigma_{LT}$ and $\sigma_{RT}$ for temperature above and below the optimum. This corresponds to $f_2(T_t^{\text{air}})$ in the main text and is written as:

$$f_2(T_t^{\text{air}}) = \exp\left[-\frac{\left(L^{j_1}(T_t^{\text{air}}) - T_{\text{opt}}\right)^2}{2\sigma_i^2}\right], \tag{1}$$

where $L^{j_1}(T_t^{\text{air}})$ indicates air temperature $j_1$ days prior, $\sigma_i = \sigma_{\text{LT}}$ when $T_t^{\text{air}} < T_{\text{opt}}$ and $\sigma_i = \sigma_{\text{RT}}$ otherwise.

The shape parameters were first fitted by minimizing the sum of squares between the normalized mechanistic EIR curve and the prediction from the simplified curve. The resulting asymmetric Gaussian curve was able to closely reproduce the more mechanistic EIR-temperature relationship (**Figure S4C**).

As part of the inference procedure described in the main text, $\sigma_{LT}$ and $\sigma_{RT}$ were estimated jointly with other parameters. The resulting temperature-EIR relationship continues to support malaria transmission above 32 degrees Celsius, as the estimated $\sigma_{RT}$ leads to a slower decline at high temperatures (**Figure S4D**).

**Figure S4:** Simplification of EIR-temperature equations. **A:** EIR. vs temperature relationship proposed in [2]. **B:** Percent of days from 2018 to 2023 whose temperature falls within a certain bin. The daily estimates come from fitting smoothing splines to average monthly temperature data as described in the main text. **C:** Finding the parameters of asymmetric Gaussian function that best match the relationship described in [2]. The values of $\sigma_{LT}$ and $\sigma_{RT}$ shown are those that minimized the squared error. Each curve was scaled so that the maximal value is one. **D:** Comparison of EIR-temperature relationship using 500 draws of $\sigma_{LT}$ and $\sigma_{RT}$ coming from the Bayesian inference procedure targeting the posterior distribution of model parameters. The solid line uses the parameters that maximized the posterior distribution and the shaded band represents the 95% credible interval

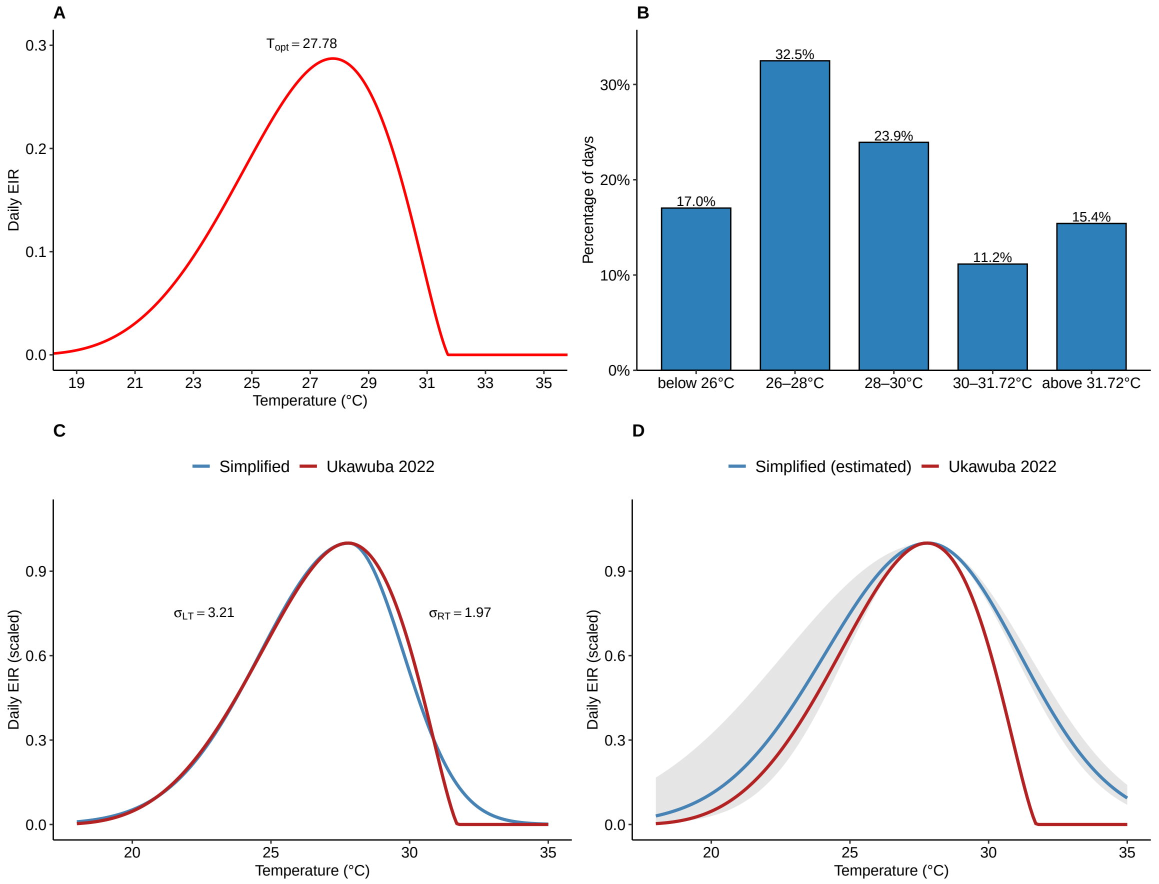

#### S2.2 Deriving birth and death rates

In order to have a well-behaved model, it was decided to have equal birth and date rates and assume that these rates are constant over the period of the analysis (2018 to 2023). In reality, estimated yearly crude death rates ranged from 14 deaths per 1000 people to 12 deaths per 1000 people and yearly estimated crude birth rates ranged from 43.2 and 47 people with a general downward trend for both indicators [3].

The exact birth and death rates were determined by first constructing the following demography model:

$$\frac{dP_1}{dt} = \delta_b(P_1 + P_2) - \delta_a P_1 - \delta_d P_1$$
$$\frac{dP_2}{dt} = \delta_a P_1 - \delta_d P_2$$

where $P_1$ and $P_2$ are the population sizes in those under 5 and 5 and older, respectively, and $\delta_a$, the daily aging rate, is equal to $1/(5 \cdot 365)$. Then, assuming $\delta_b = \delta_d$, the population sizes were simulated forward in time until equilibrium was reached, each time with a different rate. At equilibrium, the proportion of the population in $P_1$ and $P_2$ was calculated. The rate that led to the age distribution most closely matching the national estimates were chosen - 19% under 5 years old and 81% aged 5 or older [4]. This procedure resulted in a yearly crude rate of 47 deaths and births per 1000 people or, equivalently, $47/(1000 \cdot 365)$ deaths and births per person per day.

#### S2.3 Weighted likelihood via generalized Bayesian updating

To estimate the impact of SMC, the model must accurately reproduce transmission dynamics in scenarios when SMC is both present and not present. However, because SMC was deployed in five of six years, standard Bayesian inference, which minimizes a joint likelihood over all years, disproportionately emphasizes SMC-present periods. As a result, a model fitting those years well may fail to capture dynamics in 2019, the single non-SMC year.

To correct this imbalance, weights were applied to the likelihood based on the inverse frequency of each year's SMC status, following the general framework for updating prior beliefs proposed in [5]. The idea is that, while prior beliefs are generally updated through a likelihood function, beliefs can instead be updated through any generic loss function that takes as input observed data and model parameters, with the negative log-likelihood being just one case.

More formally, we let $\text{SMC}_{\text{yr}} \in \{0, 1\}$ indicate whether SMC was implemented in year yr. The weight for that year is given by:

$$\omega(\text{yr}) = \frac{p(\text{SMC} = 1)}{p(\text{SMC} = \text{SMC}_{\text{yr}})} \quad \text{where} \quad p(\text{SMC} = s) = \frac{1}{N_{\text{yr}}} \sum_{i=1}^{N_{\text{yr}}} \mathbb{1}\{\text{SMC}_i = s\}$$

The (weighted) log-likelihood function is then:

$$\ell_{\text{weighted}}(\theta, y_{w,\text{yr}}) = \omega(\text{yr}) \ell(\theta, y_{w,\text{yr}}),$$

where $\ell(\theta, y_{w,\mathrm{yr}})$ is the standard log-likelihood of a negative binomial distribution.

It is this weighted log-likelihood that was used to update the prior beliefs to construct the posterior. The end result is that the one year without SMC is up-weighted and the more frequently SMC-present years are down-weighted. Related weighting methods appear in causal inference, such as inverse probability weighting to adjust for treatment assignment imbalance [6–8], and in machine learning through class reweighting to correct for label imbalance [9].
